# Age-related differences in rejection rates, infections and tacrolimus exposure in pediatric kidney transplant recipients – a benchmark study of the CERTAIN registry

**DOI:** 10.1101/2024.05.27.24307975

**Authors:** Maral Baghai Arassi, Manuel Feißt, Kai Krupka, Atif Awan, Elisa Benetti, Ali Duzova, Isabella Guzzo, Jon Jin Kim, Sabine König, Mieczysław Litwin, Jun Oh, Anja Bücher, Lars Pape, Licia Peruzzi, Mohan Shenoy, Sara Testa, Lutz T. Weber, Jakub Zieg, Britta Höcker, Alexander Fichtner, Burkhard Tönshoff, the Cooperative European Pediatric Renal Transplant Initiative Research Network

## Abstract

**Background:** Data on age-related differences in rejection rates, infectious episodes and tacrolimus exposure in pediatric kidney transplant recipients (pKTR) on a uniform tacrolimus-based immunosuppressive regimen are scarce.

**Methods:** We therefore performed a large-scale analysis of 802 pKTR from the CERTAIN registry from 40 centers in 14 countries. Inclusion criteria were a tacrolimus-based immunosuppressive regimen and at least two years of follow-up. The patient population was divided into three age groups (infants <6 years, school-aged children 6-12 years, and adolescents >12 years) to assess age-related differences in outcome.

**Results:** Median follow-up was 48 months (IQR, 36-72). Within the first 2 years post-transplant, infants had a significantly higher incidence of infections (80.6% vs. 55.0% in adolescents, P<0.001) and a significantly higher number of cumulative hospital days (median 13 days vs. 7 days in adolescents, P < 0.001). Adolescents had a significantly higher rate of biopsy-proven acute rejection episodes in the first year post-transplant (21.7%) than infants (12.6%, P=0.007). Infants had significantly lower tacrolimus trough levels, lower concentration-to-dose ratios as an approximation for higher tacrolimus clearance, and higher intra-patient variability (all P < 0.01) than adolescents.

**Conclusions:** This largest study to date in European pKTR on a tacrolimus-based immunosuppressive regimen shows important age-related differences in rejection rates, infection episodes, tacrolimus exposure and clearance. These data suggest that immunosuppressive therapy in pKTR should be tailored according to the age-specific risk profiles of this heterogeneous patient population.

## Introduction

Kidney transplantation is the preferred option for the treatment of kidney failure. However, allograft rejection, infection, and graft dysfunction challenge patient and graft survival. While large multicenter outcomes studies exist in adult transplant cohorts [1, 2], there is a paucity of similar studies in the field of pediatric kidney transplantation, because robust and generalizable outcomes studies in pediatric cohorts are difficult to achieve due to the small number of pediatric patients requiring kidney transplantation. However, there is an urgent need for large, multicenter studies that analyze age-related differences in outcomes in this heterogeneous patient population. These data may support clinical decision making and thereby improve patient care.

Benchmarks play a key role in filling this data gap. They provide a framework for assessing the effectiveness of treatment protocols and help to evaluate and improve current practices. While benchmark studies of North American and Chinese pediatric cohorts are available [3, 4], similar studies are lacking in Europe. The *Cooperative European Paediatric Renal Transplant Initiative* (CERTAIN) registry provides a suitable tool to address this need. It currently includes 95 pediatric kidney transplant centers from 26 European countries and data from 3930 pediatric patients. CERTAIN is a platform for conducting high quality research due to its detailed records and thorough follow-up. It is equipped to analyze long-term outcomes and trends in pediatric kidney transplantation.

In this study, we used the CERTAIN registry to fill the gap in age-related outcome data for pediatric kidney transplant recipients treated with a tacrolimus-based immunosuppressive regimen. Tacrolimus was chosen because of its widespread use and its status as the leading immunosuppressive agent, a position it is expected to maintain for the next decade. This choice also allows for a more uniform patient cohort, while still representing the vast majority of pediatric kidney transplant recipients. Our analysis included data from 802 children at 40 centers in 14 countries over a 20-year period. This research provides critical insight into the impact of age on kidney transplant outcomes and highlights the unique challenges faced by each age group.

## Patients and Methods

### Study Design and Patient Population

This is a retrospective, multicenter, longitudinal cohort study using data from the CERTAIN registry, including patients who underwent kidney transplantation between February 1999 and April 2019. The CERTAIN registry collects detailed longitudinal clinical and laboratory data and applies rigorous validity checking procedures (http://www.certain-registry.eu/). Participation in the CERTAIN registry is approved by the ethics committee at each center. Informed consent was obtained from the parents or legal guardians prior to enrollment, with assent from patients when appropriate for their age. The time points of data collection and the corresponding time intervals were as follows: baseline (pre-transplant), at months 1, 3, 6, 9, 12, and every 6 months thereafter (see Supplementary Methods for detailed description of the CERTAIN registry). All procedures and immunosuppressive regimens were performed according to local institutional protocols. Anthropometric, clinical, and biochemical data were collected as part of a routine follow-up at each center. The study was conducted in accordance with the Declaration of Helsinki and the Declaration of Istanbul on Organ Trafficking and Transplant Tourism. The study was designed, analyzed, and reported according to the STROBE guidelines (https://www.strobe-statement.org/).

### Inclusion/Exclusion Criteria

The study included kidney allograft recipients who were 21 years of age or younger at the time of transplantation and were maintained on a tacrolimus-based immunosuppressive regimen throughout the follow-up period. Eligibility required a complete data set for at least two years post-transplant. Patients with ABO-incompatible transplants or those who received additional non-kidney allografts were excluded. Figure 1 provides an overview of the patient inclusion and exclusion criteria.

**Fig. 1:**
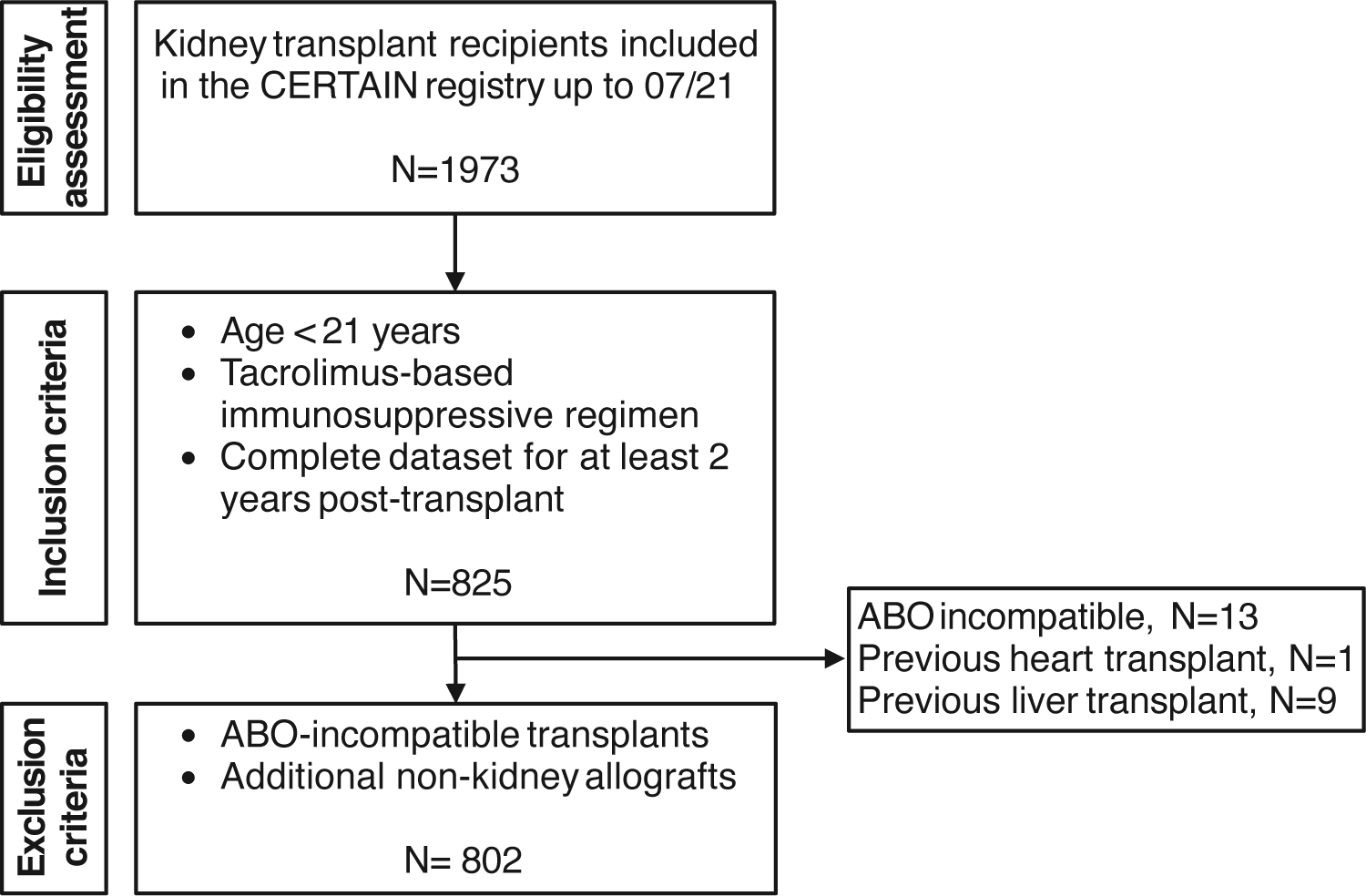
Flow diagram of patient inclusion and exclusion. Patient flow diagram showing the inclusion and exclusion criteria applied to the CERTAIN registry data to generate the cohort for the current study.

### Outcome events

Study outcomes included infection, rejection, graft dysfunction, diabetes mellitus, death, and cumulative hospital days. Because graft loss is a rare event in pediatric KTx recipients, the outcome measure we used was a death-censored composite end point named allograft dysfunction, which is defined as either graft loss, or eGFR ≤30 mL/min per 1.73 m², or a ≥50% decline from baseline eGFR at month 3 post-transplant. eGFR values above 120 mL/min per 1.73 m² at month 3 post-transplant were set to 120 mL/min per 1.73m². Rejections were classified as such if they were confirmed by biopsy and received antirejection therapy. They were further categorized based on graft histopathology using the most recent Banff classification in effect at the time of rejection (1997-2019). Infections were identified based on the treating physician’s determination of an episode of infection, using a binary survey of infection without any detailed specification. For cytomegalovirus (CMV), data on organ involvement and CMV disease were also available. Episodes of rejection and infection were categorized according to their timing, distinguishing between those occurring in the 1^st^ or 2^nd^ year post-transplant. Recurrent infections and rejections were defined as > 1 event during follow-up.

### Tacrolimus exposure quantification

Tacrolimus exposure was quantified using whole blood trough levels as documented in the CERTAIN database. Trough level data were categorized into early (months 1-3), mid (months 6-12), and late (months 18-24) post-transplant periods. To assess tacrolimus intrapatient variability (TacIPV), we required a minimum of three tacrolimus trough level measurements per patient during the 6-12 month post-transplant period. TacIPV was assessed using two algorithms: the observed coefficient of variation (CV_obs_), calculated as (standard deviation ÷ mean) * 100 and expressed as a percentage [5, 6], and the observed mean absolute deviation (MAD_obs_), formulated as {[(Xmean-X1) + (Xmean-X2) + … + (Xmean-Xn)] ÷ n} ÷ Xmean, where X is the tacrolimus trough level in whole blood [5, 7, 8]. In addition, the concentration-to-dose (C/D) ratio was determined from tacrolimus trough levels and the corresponding daily doses. C/D ratio data were stratified by early (mean C/D ratio of months 1 and 3), late (mean C/D ratio of months 6, 9, and 12), and total (mean C/D ratio of months 1 to 12) post-transplant periods. To account for differences in body size in this pediatric patient population, C/D ratio data were corrected for body surface area (C/D ratio_BSA_ = Tac blood trough level [ng/ml] ÷ (daily Tac dose [mg]/body surface area [m²])) and for body weight (C/D ratio_BW_ = Tac blood trough level [ng/ml] ÷ (daily Tac dose [mg]/body weight [kg])). To increase comparability with adult C/D ratios, BSA-corrected C/D ratio was additionally adjusted for a representative adult BSA of 1.73 m^2^ (C/D ratio_BSA-Adult_ = Tac blood trough level [ng/ml] ÷ (daily Tac dose [mg]/body surface area [m²]*1.73).

### Statistical analysis

Continuous variables were described by mean and standard deviation or median and interquartile range, as appropriate, and categorical variables were described by absolute and relative frequencies. Comparisons between the three age subgroups were made using F-tests for continuous variables and chi-squared for categorical variables. Post hoc tests were corrected using the Benjamini-Hochberg correction. Time-to-event variables were described using the Kaplan-Meier estimator, and comparisons were made using log-rank tests. Univariable and multivariable regression models were used to identify risk factors. Linear regression models were used for cumulative hospital days (in relation to follow-up time), and Cox regression analysis was used for time-to-event outcomes. Variables with P values less than 0.1 in the subgroup analyses and univariable analyses were included in the multivariable analyses after medical plausibility screening. Model estimates were reported with corresponding 95% confidence intervals for each measure (i.e., hazard ratios, inverse hazard ratios, regression estimates). To assess the influence of a potential era effect on patient outcomes while maintaining an even distribution of patient numbers across groups, data were categorized into two different time periods of kidney transplantation: 08/1999-07/2014 (era 1, N=402) and 08/2014-04/2019 (era 2, N=400). All analyses were performed in R version >4.2.0. Missing values were very rare and no imputation of missing values was performed. In this exploratory analysis, all P values should be interpreted in a descriptive sense without a confirmatory value.

## Results

### Patient characteristics

A total of 802 patients, 323 girls (40.3%) and 479 boys (59.7%), who underwent kidney transplantation between 1999 and 2019 from 40 different study centers were included (Table 1, Table S1). The median age at transplantation was 11.1 (IQR, 6.85-15.3) years. The majority of patients were of Caucasian descent (91.4%), with a minority of patients of African (3.1%) or Asian (5.4%) descent. In the majority of patients, the primary kidney disease was congenital anomalies of the kidney and urinary tract (CAKUT, 39.4%), followed by hereditary cystic or congenital diseases (25.3%) and primary glomerular diseases (17.6%). Sixty-nine percent of patients received a deceased donor kidney. All patients received tacrolimus, MMF, and a glucocorticoid as initial immunosuppressive regimen. Forty-one percent of patients received additional induction therapy with either basiliximab (34.2%), anti-thymocyte globulin (5.4%), rituximab (2.0%), or daclizumab (0.7%). The median follow-up time was 48 (IQR, 36-72) months post-transplant. The patient cohort was stratified into three age groups: < 6 years (N=206, infants), 6-12 years, (N=278, school-aged children), > 12 years (N=318, adolescents). Age groups differed significantly with respect to sex, follow-up time, preemptive transplantation, primary kidney disease, induction therapy, previous kidney transplants, immunosuppressive therapy at year 1 and tacrolimus formulation. Baseline clinical and demographic characteristics are shown in Table 1.

**Table 1:**
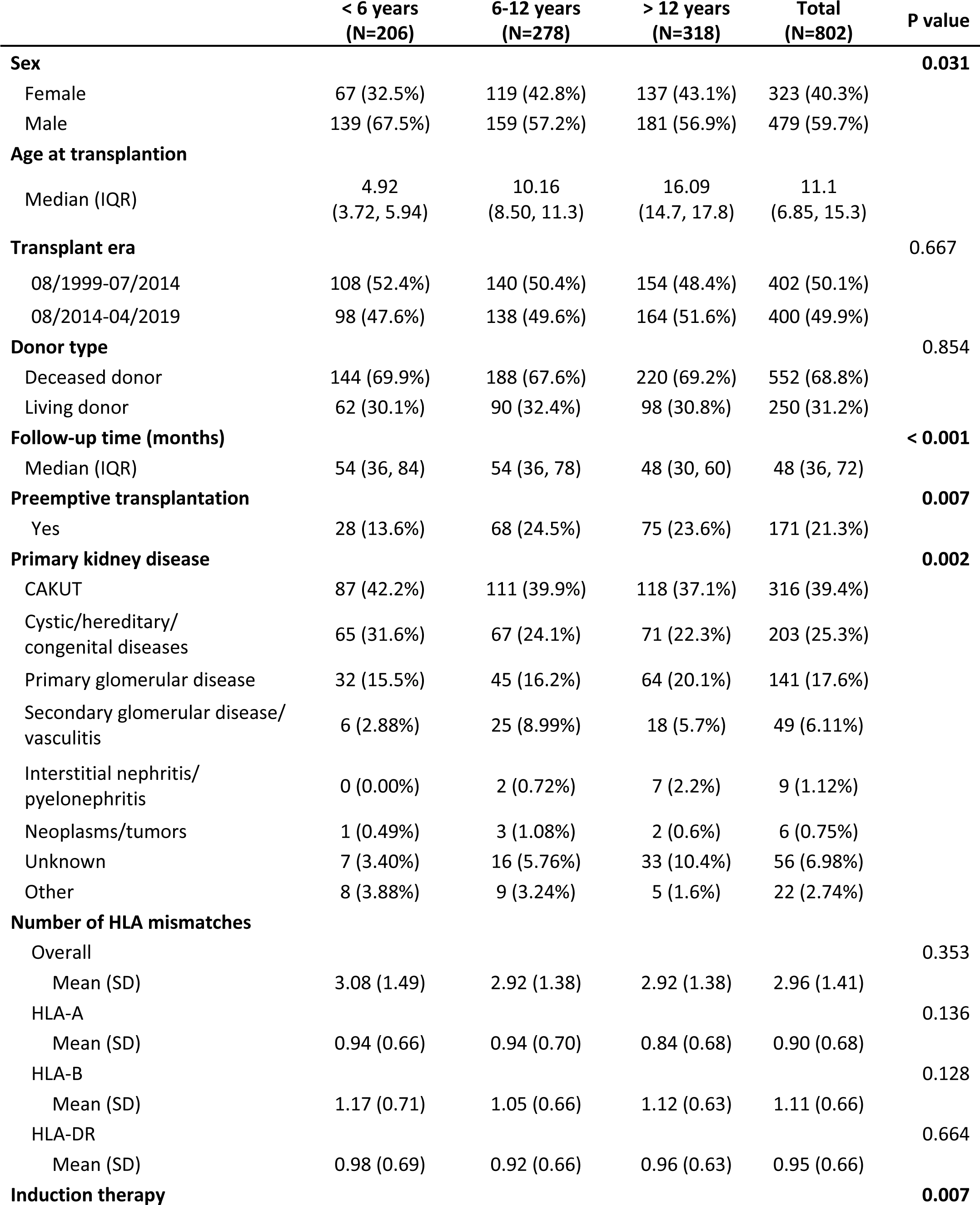
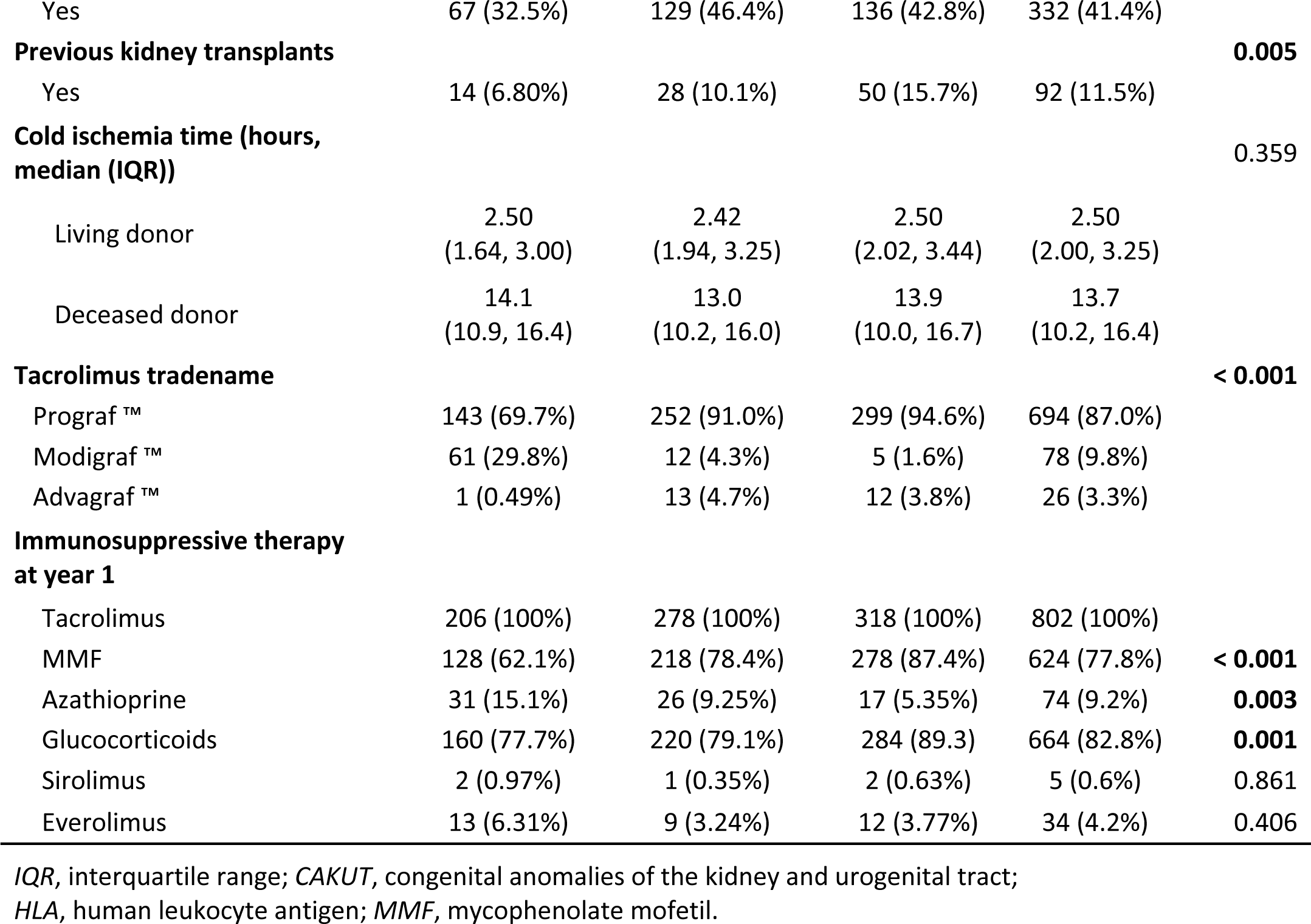
Baseline clinical and demographic characteristics.

|  | < 6 years<br>(N=206) | 6-12 years<br>(N=278) | > 12 years<br>(N=318) | Total<br>(N=802) | P value |
| --- | --- | --- | --- | --- | --- |
| **Sex** |  |  |  |  | **0.031** |
| Female | 67 (32.5%) | 119 (42.8%) | 137 (43.1%) | 323 (40.3%) |  |
| Male | 139 (67.5%) | 159 (57.2%) | 181 (56.9%) | 479 (59.7%) |  |
| **Age at transplantation** |  |  |  |  |  |
| Median (IQR) | 4.92<br>(3.72, 5.94) | 10.16<br>(8.50, 11.3) | 16.09<br>(14.7, 17.8) | 11.1<br>(6.85, 15.3) |  |
| **Transplant era** |  |  |  |  | 0.667 |
| 08/1999-07/2014 | 108 (52.4%) | 140 (50.4%) | 154 (48.4%) | 402 (50.1%) |  |
| 08/2014-04/2019 | 98 (47.6%) | 138 (49.6%) | 164 (51.6%) | 400 (49.9%) |  |
| **Donor type** |  |  |  |  | 0.854 |
| Deceased donor | 144 (69.9%) | 188 (67.6%) | 220 (69.2%) | 552 (68.8%) |  |
| Living donor | 62 (30.1%) | 90 (32.4%) | 98 (30.8%) | 250 (31.2%) |  |
| **Follow-up time (months)** |  |  |  |  | < 0.001 |
| Median (IQR) | 54 (36, 84) | 54 (36, 78) | 48 (30, 60) | 48 (36, 72) |  |
| **Preemptive transplantation** |  |  |  |  | **0.007** |
| Yes | 28 (13.6%) | 68 (24.5%) | 75 (23.6%) | 171 (21.3%) |  |
| **Primary kidney disease** |  |  |  |  | **0.002** |
| CAKUT | 87 (42.2%) | 111 (39.9%) | 118 (37.1%) | 316 (39.4%) |  |
| Cystic/hereditary/<br>congenital diseases | 65 (31.6%) | 67 (24.1%) | 71 (22.3%) | 203 (25.3%) |  |
| Primary glomerular disease | 32 (15.5%) | 45 (16.2%) | 64 (20.1%) | 141 (17.6%) |  |
| Secondary glomerular disease/<br>vasculitis | 6 (2.88%) | 25 (8.99%) | 18 (5.7%) | 49 (6.11%) |  |
| Interstitial nephritis/<br>pyelonephritis | 0 (0.00%) | 2 (0.72%) | 7 (2.2%) | 9 (1.12%) |  |
| Neoplasms/tumors | 1 (0.49%) | 3 (1.08%) | 2 (0.6%) | 6 (0.75%) |  |
| Unknown | 7 (3.40%) | 16 (5.76%) | 33 (10.4%) | 56 (6.98%) |  |
| Other | 8 (3.88%) | 9 (3.24%) | 5 (1.6%) | 22 (2.74%) |  |
| **Number of HLA mismatches** |  |  |  |  |  |
| Overall |  |  |  |  | 0.353 |
| Mean (SD) | 3.08 (1.49) | 2.92 (1.38) | 2.92 (1.38) | 2.96 (1.41) |  |
| HLA-A |  |  |  |  | 0.136 |
| Mean (SD) | 0.94 (0.66) | 0.94 (0.70) | 0.84 (0.68) | 0.90 (0.68) |  |
| HLA-B |  |  |  |  | 0.128 |
| Mean (SD) | 1.17 (0.71) | 1.05 (0.66) | 1.12 (0.63) | 1.11 (0.66) |  |
| HLA-DR |  |  |  |  | 0.664 |
| Mean (SD) | 0.98 (0.69) | 0.92 (0.66) | 0.96 (0.63) | 0.95 (0.66) |  |
| **Induction therapy** |  |  |  |  | **0.007** |
| Yes | 67 (32.5%) | 129 (46.4%) | 136 (42.8%) | 332 (41.4%) |  |
| **Previous kidney transplants** |  |  |  |  | **0.005** |
| Yes | 14 (6.80%) | 28 (10.1%) | 50 (15.7%) | 92 (11.5%) |  |
| **Cold ischemia time (hours, median (IQR))** |  |  |  |  | **0.359** |
| Living donor | 2.50<br>(1.64, 3.00) | 2.42<br>(1.94, 3.25) | 2.50<br>(2.02, 3.44) | 2.50<br>(2.00, 3.25) |  |
| Deceased donor | 14.1<br>(10.9, 16.4) | 13.0<br>(10.2, 16.0) | 13.9<br>(10.0, 16.7) | 13.7<br>(10.2, 16.4) |  |
| **Tacrolimus tradename** |  |  |  |  | **< 0.001** |
| Prograf <sup>™</sup> | 143 (69.7%) | 252 (91.0%) | 299 (94.6%) | 694 (87.0%) |  |
| Modigraf <sup>™</sup> | 61 (29.8%) | 12 (4.3%) | 5 (1.6%) | 78 (9.8%) |  |
| Advagraf <sup>™</sup> | 1 (0.49%) | 13 (4.7%) | 12 (3.8%) | 26 (3.3%) |  |
| **Immunosuppressive therapy at year 1** |  |  |  |  |  |
| Tacrolimus | 206 (100%) | 278 (100%) | 318 (100%) | 802 (100%) |  |
| MMF | 128 (62.1%) | 218 (78.4%) | 278 (87.4%) | 624 (77.8%) | **< 0.001** |
| Azathioprine | 31 (15.1%) | 26 (9.25%) | 17 (5.35%) | 74 (9.2%) | **0.003** |
| Glucocorticoids | 160 (77.7%) | 220 (79.1%) | 284 (89.3) | 664 (82.8%) | **0.001** |
| Sirolimus | 2 (0.97%) | 1 (0.35%) | 2 (0.63%) | 5 (0.6%) | 0.861 |
| Everolimus | 13 (6.31%) | 9 (3.24%) | 12 (3.77%) | 34 (4.2%) | 0.406 |

*IQR*, interquartile range; *CAKUT*, congenital anomalies of the kidney and urogenital tract; *HLA*, human leukocyte antigen; *MMF*, mycophenolate mofetil.

### Biopsy-proven rejections

Biopsy-proven rejection episodes were the second most common event during the first 2 years post-transplant, occurring at least once in 23.8% of patients. The prevalence of rejection differed significantly by age, with the highest rate in adolescents (28.9% vs. 18.9% in infants, P=0.018). When stratified by year, most rejections occurred in the first year post-transplant (Figure 2a), with adolescents experiencing significantly (P=0.035) more rejections than infants and school-aged children (P=0.043). Notably, adolescents experienced also a higher rate of recurrent rejections (10.4%) than school-aged children (4.7%) or infants (4.9%, P=0.042). Also in the Kaplan-Meier analysis, the rejection-free survival was significantly (P=0.027) higher in infants than in adolescents throughout the entire follow-up period (Figure 3). This trend was maintained (P=0.068), when patients with a history of more than one kidney transplant were excluded (N=92, Table 1, Figure S1). Rejection episodes were further categorized based on graft histopathology using the most recent Banff classification in effect at the time of rejection (1997-2019, Figure 2b). Table S2 provides an overview of rejection subtypes stratified by age group and years 1 and 2 post-transplant. Borderline rejection was the most common finding, followed by T cell-mediated rejection (TCMR); the latter was significantly more frequent in adolescents (P=0.015) than in infants (Figure 2b). Multivariable Cox regression analysis revealed an increased risk of rejection in adolescents (HR, 1.53; 95% CI, 1.10-2.13; P=0.01). Overall rejection rates were comparable between transplant era 1 and 2.

**Fig. 2:**
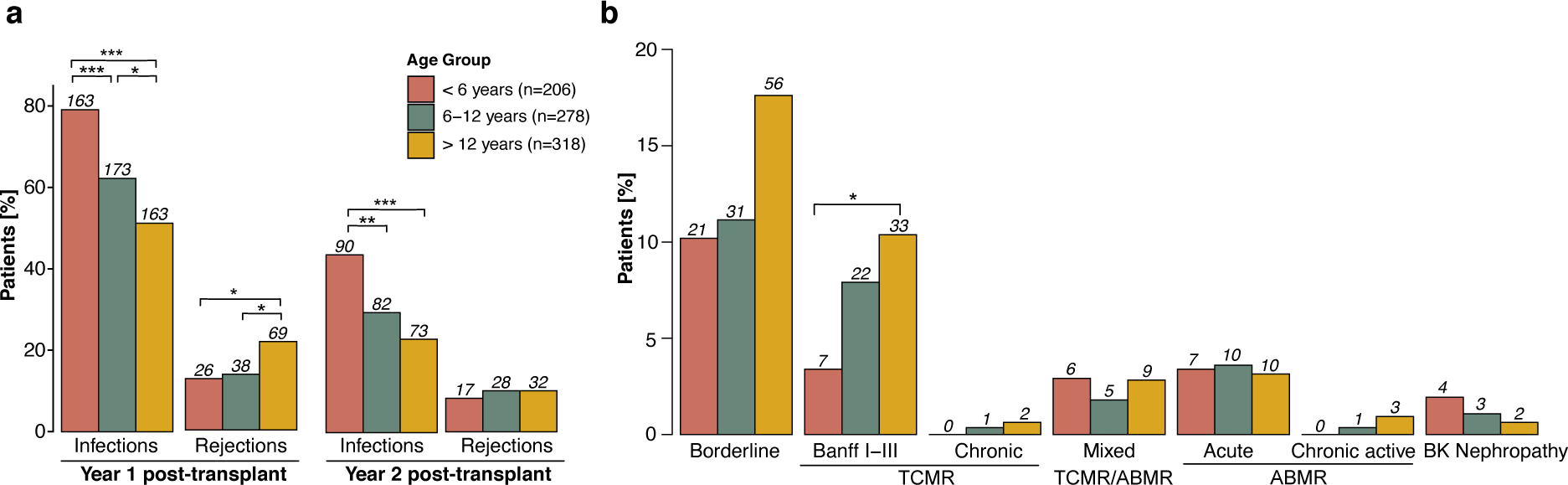
Age-stratified outcome parameters in the first two years post-transplant. **(a)** Incidence of first episodes of rejection and infection in the first two years post-transplant, stratified by age group and year post-transplant. The first event of each patient in each year is considered for analysis. **(b)** Histopathology of allograft rejection graded according to Banff in the first two years post-transplant, stratified by age group. Color coding represents different age groups. For detailed rejection subtypes stratified by year see Table S2. Benjamini-Hochberg-corrected statistical significance values are indicated by asterisks as follows: *P < 0.05, **P < 0.01, ***P < 0.001. ABMR, antibody-mediated rejection; BK, BK polyomavirus; TCMR, T cell-mediated rejection.

**Fig. 3:**
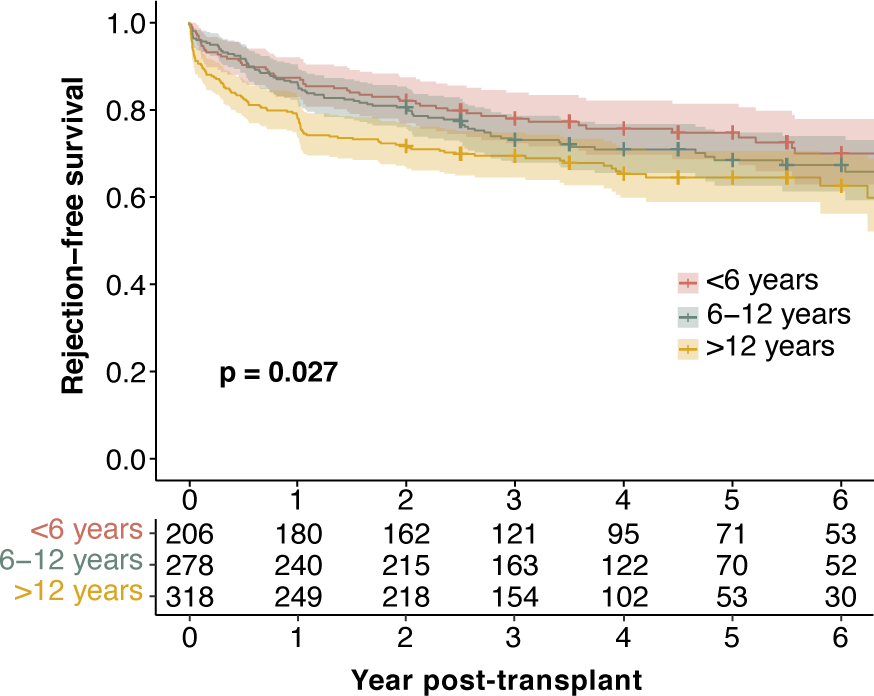
Rejection-free survival stratified by age group. Rejection-free survival: Kaplan-Meier curves illustrating the probability of rejection-free survival over the entire follow-up period post-transplant, with age groups distinguished by color coding. Log-rank P values from the Kaplan-Meier analysis are provided to indicate the statistical significance of differences between age groups.

### Infections

During the first two years post-transplant, infections were the most common outcome event, with 65.2% of patients experiencing at least one infectious episode. Infants had the highest incidence of infections (80.6%), followed by school-aged children (65.5%) and adolescents (55.0%, P<0.001, Figure 2A). Infants also had the highest incidence of recurrent infections (66.5% vs. 33.6% in adolescents, P <0.001). This pattern was consistent for both the 1-year and the 2-year post-transplant periods (both P<0.001). Infection rates were comparable between transplant era 1 and 2. When categorizing infections by type (Fig. 4, Table S3), gastroenteritis was the most common, closely followed by lower respiratory tract infections and pyelonephritis (all P<0.001 for infants vs. adolescents). Notably, infants experienced more severe infections such as sepsis (P=0.011) and other relevant infections such as EBV (P<0.001) and BKPyV (P=0.002). CMV infections did not differ significantly between age groups. In the majority of events (92% in infants and school-aged children, 83% in adolescents), CMV infection was only CMV viremia as detected by nuclear acid testing or pp65 antigenemia, without CMV syndrome or organ-invasive disease. Multivariable Cox regression analysis showed a significantly higher risk of infection in infants (inverse hazard ratio, 1.85; 95% CI, 1.51-2.33; P<0.001) than in adolescents (Table 2) and school-aged children (inverse hazard ratio, 1.39; 95% CI, 1.12-1.69; P=0.002).

**Fig. 4:**
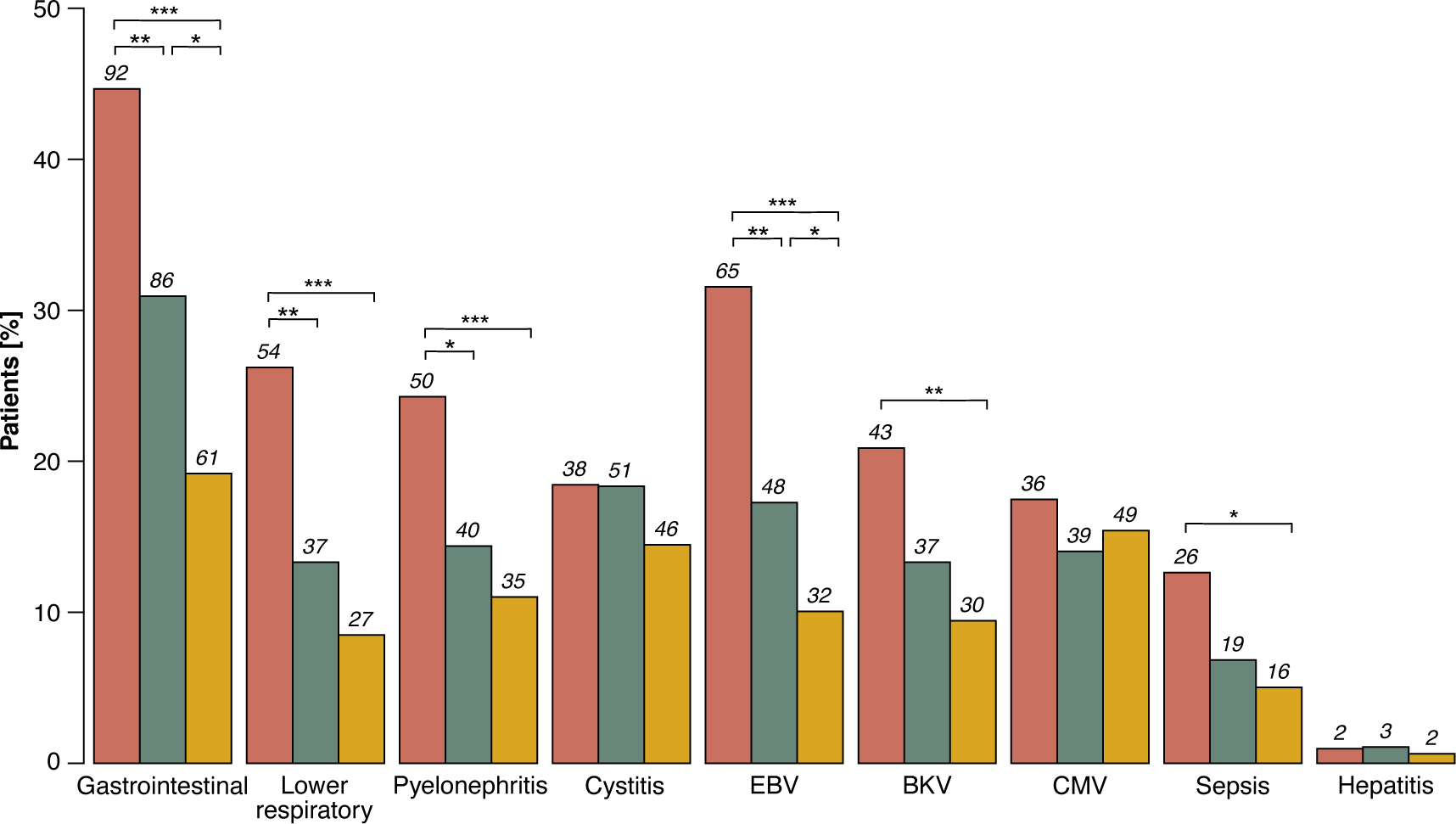
Types of infections observed within the first two years post-transplant. Distribution of infection types observed within the first two years post-transplant, stratified by age group. For detailed infection subtypes in each category see Table S3. Color coding represents different age groups. Benjamini-Hochberg-corrected statistical significance values are indicated by asterisks as follows: *P < 0.05, **P < 0.01, ***P < 0.001. BKV, BK pyeloma virus; CMV, cytomegalovirus; EBV, Epstein-Barr virus.

**Table 2:** Risk factor analysis for outcome events.

|  | Unadjusted HR/<br>Estimate (95% CI) | P<br>value | Adjusted HR/<br>Estimate (95% CI) | P<br>value |
| --- | --- | --- | --- | --- |
| **Infections** |  |  |  |  |
| Age group 6-12 years | 0.73 (0.6-0.9) | **0.002** | 0.72 (0.59-0.89) | **0.002** |
| Age group > 12 years | 0.54 (0.44-0.66) | **<0.001** | 0.54 (0.43-0.66) | **<0.001** |
| Male sex | 1.09 (0.93-1.29) | 0.29 | 1.04 (0.88-1.24) | 0.65 |
| Preemptive transplantation | 0.9 (0.74-1.1) | 0.31 | 0.90 (0.73-1.01) | 0.33 |
| CAKUT | 1.12 (0.94-1.30) | 0.22 | 1.09 (0.92-1.30) | 0.3 |
| HLA-Mismatches | 0.95 (0.90-1.10) | 0.09 | 0.94 (0.89-1.00) | 0.04 |
| Induction therapy | 0.98 (0.83-1.15) | 0.78 | 1.03 (0.88-1.22) | 0.7 |
| **Rejections** |  |  |  |  |
| Age group 6-12 years | 1.162 (0.83-1.63) | 0.383 | 1.15 (0.81-1.62) | 0.43 |
| Age group > 12 years | 1.47 (1.05-2.05) | **0.012** | 1.53 (1.10-2.13) | **0.01** |
| Male sex | 1.13 (0.88-1.46) | 0.17 | 1.22 (0.94-1.59) | 0.14 |
| Preemptive transplantation | 1.08 (0.8-1.45) | 0.63 | 1.04 (0.77-1.41) | 0.79 |
| CAKUT | 0.92 (0.71-1.19) | 0.51 | 0.87 (0.66-1.13) | 0.29 |
| HLA-Mismatches | 1.01 (0.92-1.10) | 0.88 | 1.01 (0.77-1.41) | 0.79 |
| Induction therapy | 1.2 (0.94-1.54) | 0.15 | 1.15 (0.9-1.49) | 0.27 |
| **Hospitalization days** |  |  |  |  |
| Age group 6-12 years | -2.17 (-3.85- -0.49) | **0.01** | -2.12 (-3.85- -0.39) | **0.02** |
| Age group > 12 years | -2.5 (-4.13- -0.87)) | **0.003** | -2.41 (-4.09- -0.73)) | **0.005** |
| Male sex | 0.80 (-0.52-2.12) | 0.23 | 0.68 (-0.70-2.07) | 0.33 |
| Preemptive transplantation | -1.34 (-2.92-0.24) | 0.1 | -1.04 (-2.68-0.6) | 0.21 |
| CAKUT | 0.46 (-0.87-1.79) | 0.5 | 0.23 (-1.17-1.63) | 0.74 |
| HLA-Mismatches | 0.345 (-0.12-0.81) | 0.14 | 0.28 (-0.19-0.74) | 0.25 |
| Induction therapy | 0.35 (-0.97-1.67) | 0.6 | 0.37 (-0.97-1.7) | 0.59 |

*HR*, hazard ratio; *CI*, confidence interval, CAKUT, congenital anomalies of the kidney and urogenital tract; HLA, human leukocyte antigen

### Graft dysfunction, diabetes mellitus, death

The overall incidence of graft dysfunction (2.8%), diabetes mellitus (1.7%), and death (0.1%) during the first 2 years post-transplant was low. Diabetes mellitus was significantly more common in adolescents (3.8%) than in school-aged children (0.7%) and infants (0%; P=0.002). Kaplan-Meier analysis over the entire follow-up period also showed no significant age differences for graft dysfunction or death.

### Hospitalizations

Ninety-one percent of patients experienced at least one hospitalization during the first two years post-transplant, with no significant difference between age groups (P=0.973). The most common reason for hospitalization was infection, which was progressively more common in infants and school-aged children than in adolescents (P<0.001, Figure 5). In addition, the total number of days spent in the hospital during the first 2 years post-transplant was significantly (P<0.001) higher in infants (median 13 days, IQR 4-33) than in adolescents (median 7 days, IQR 2-18) and school-aged children (median 7 days, IQR 2-21). Cumulative hospitalization days were comparable between transplant era 1 and 2. Rejection episodes were a more frequent cause of hospitalization in adolescents than in infants (P<0.001) and school-aged children (P=0.001), even when patients when multiple kidney transplantations were excluded (P<0.001 and P=0.001). Multivariable linear regression analysis showed that infants had a significantly higher risk of hospitalization (estimate of 2.41, 95% CI, 0.73-4.09, P=0.005) than adolescents (Table 2) and than school-aged children (2.12, 95% CI, 0.39-3.85, P=0.02).

**Fig. 5:**
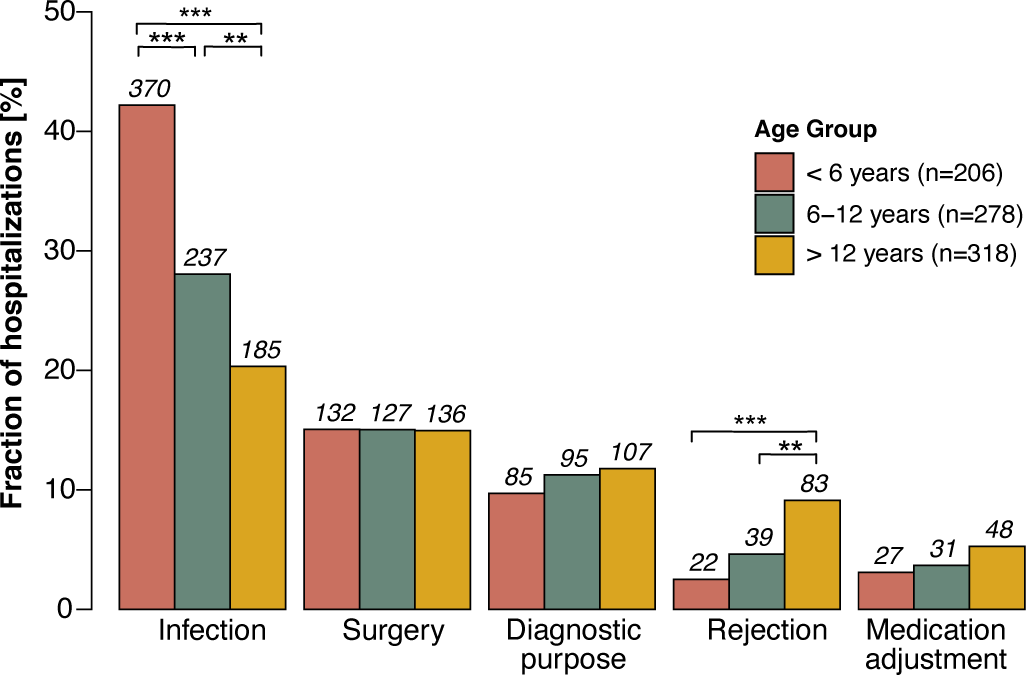
Reasons for hospitalization in the first two years post-transplant. Distribution of reasons for hospitalization in the first two years post-transplant, stratified by age group. Color coding represents different age groups. Benjamini-Hochberg-corrected statistical significance values are indicated by asterisks as follows: *P < 0.05, **P < 0.01, ***P < 0.001.

### Quantification of tacrolimus exposure

Mean tacrolimus trough levels ranged from 8.50 ± 2.13 ng/mL at month 1-3 post-transplant to 5.95 ± 1.62 ng/mL at months 18-24 post-transplant (Fig. 6a). In all investigated post-transplant periods (early (months 1-3), mid (months 6-12), and late (months 18-24)), infants had lower tacrolimus trough levels than school-aged children, who in turn had lower tacrolimus trough levels than adolescents (Figure 6a). These differences remained significant (P<0.05) when patients with previous kidney transplantations were excluded. For the entire cohort, the mean TacIPV expressed as CV was 18.9% ± 14.7% and expressed as MAD was 13.9% ± 11.4%. For both TacIPV algorithms, adolescents had significantly lower TacIPV values than school-aged children (P< 0.03) and infants (P=0.002, Figure 6b). The mean C/D ratio corrected for body surface area was 1.71 ± 1.01 (Figure 6c), 49.0 ± 35.7 corrected for body weight (Figure 6d), and 1.02 ± 0.68 corrected for body surface area adjusted to 1.73 m^2^ (Figure 6e). The respective age-specific C/D ratios were not significantly different between the month 1-3 and month 6-12 post-transplant periods. For all three C/D ratio algorithms, infants had significantly lower values than school-aged children, who in turn had lower values than adolescents (Figure 6c-e). None of the tacrolimus exposure parameters were associated with any of the outcome events.

**Fig. 6:**
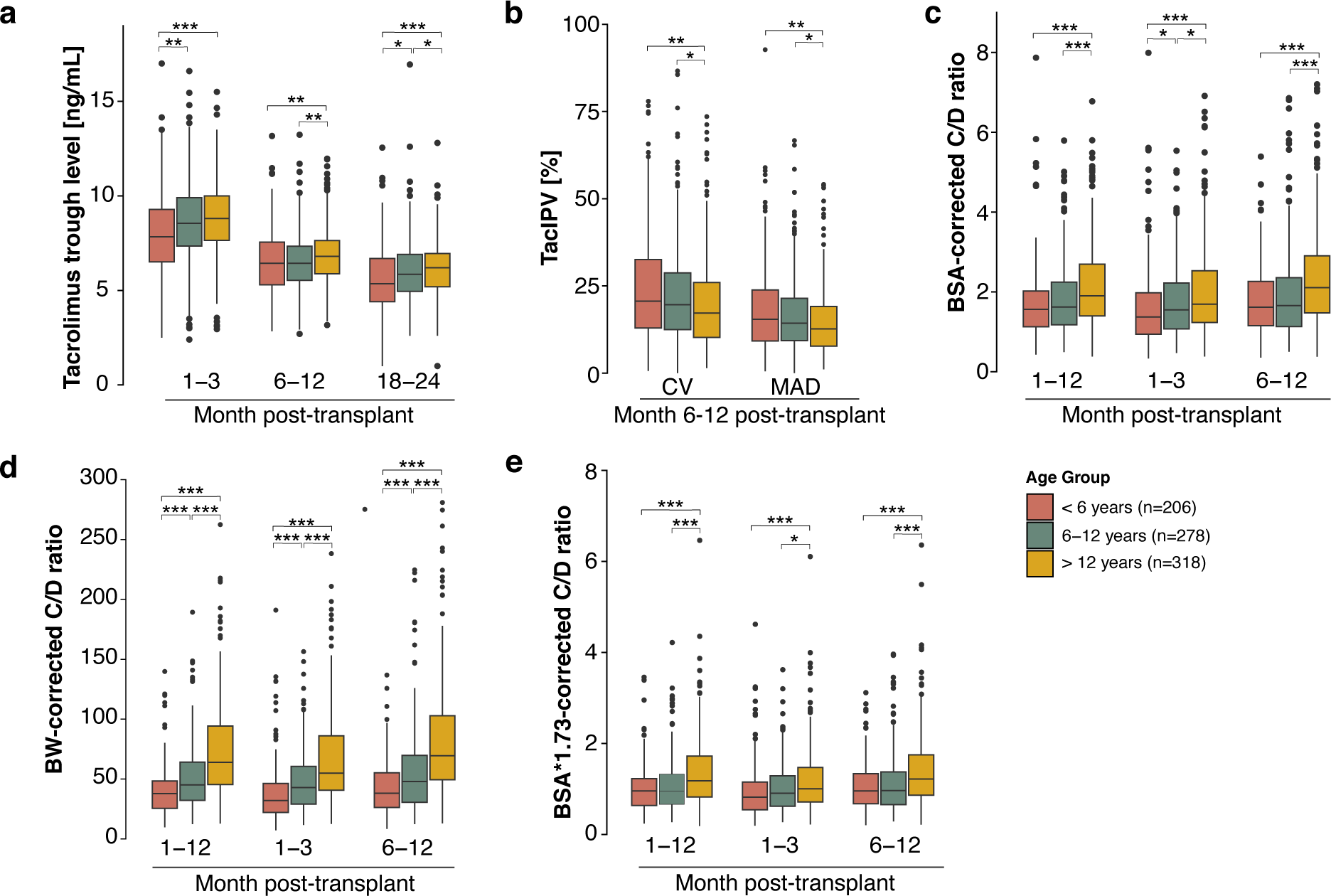
Tacrolimus exposure parameters stratified by age group. **(a)** Tacrolimus trough levels, **(b)** tacrolimus intrapatient variability (TacIPV) quantified by coefficient of variation (CV) and mean absolute deviation (MAD), **(c)** Concentration/dose ratio (C/D ratio) corrected for body surface area (BSA), **(d)** C/D ratio corrected for body weight (BW), **(e)** C/D ratio corrected for BSA scaled for 1.73 m^2^ stratified by age and time period post-transplant. Color coding represents different age groups. Benjamini-Hochberg-corrected statistical significance values are indicated by asterisks as follows: *P < 0.05, **P < 0.01, ***P < 0.001.

## Discussion

This is the largest study to date on age-related differences in rejection rates, infections and tacrolimus exposure in pediatric kidney transplant recipients on a tacrolimus-based immunosuppressive regimen. Infants had higher rates of relevant infections and more hospital days, but lower rejection rates, suggesting the need to explore less intensive immunosuppressive strategies to balance infection and rejection risks.

Our study uniquely highlights how adolescents experienced higher rates of rejection and associated hospitalizations. This pattern was statistically significant only during the first year post-transplant and in the Kaplan-Meier analysis over the entire follow-up period. While a similar pattern was observed during the second year post-transplant (Figure 2a) and after exclusion of patients with previous kidney transplants, it did not reach statistical significance (P=0.068, Figure S1). These data suggest that age and its associated psychosocial characteristics such as non-adherence may be a significant determinant of the rejection risk in pediatric kidney transplant recipients at least during the first 2 years post-transplant, but that the higher rejection risk in adolescents is also due to the higher percentage (15.7%) of patients with previous kidney transplants which may have led to sensitization to HLA antigens. Since patient enrollment began in 1999, we observed no significant era effects on rejection, infection and hospitalization rates, which is consistent with recent NAPRTCS findings showing minimal impact of era effects on outcomes between 1997 and 2007 [4].

Our data show that younger patients have lower tacrolimus trough levels, and higher TacIPV. This contrasts with other studies suggesting higher TacIPV in adolescents due to lower adherence to the immunosuppressive medication [9–11], but is consistent with our previous single-center study [12] and previous work by Prytula et al. [13] showing higher TacIPV in younger patients. Although our study does not allow for causal inferences, potential factors leading to higher tacrolimus variability could be differences in metabolism [14], frequency of dose adjustments due to intercurrent infections [5, 15], especially diarrhea which is more common in infants and may lead to a paradoxical increase in tacrolimus exposure, infection-related inflammation [16], and gut microbiota composition between young children and adolescents [17, 18]. Our study is also the first to report on age-related differences of the tacrolimus C/D ratio in pediatric kidney transplant recipients corrected for BSA and adjusted BSA to 1.73 m^2^, allowing for comparability of different C/D ratio algorithms and comparability with adult C/D ratio values. The lower C/D ratio in younger patients is consistent with the concept of a higher tacrolimus clearance in this age group, which may be due to a higher liver weight/body weight ratio in younger patients [19] and/or age-related differences in CYP3A4 and/or CYP3A5 enzyme activity [19]. This is also consistent with the lower BSA-corrected and BSA-adjusted for 1.73m^2^-corrected C/D ratios in our cohort compared to adult cohorts, although comparability is limited due to differences in the time period of C/D ratio quantification and concomitant immunosuppression [20, 21]. In contrast to a study by Naesens et al. on BW-corrected C/D ratios in pediatric kidney transplant recipients, our values appear to be lower (120 ± 93.9 at month 6 post-transplant vs. 52.9 ± 42.8 during months 6-12 post-transplant) [22]. This difference could be due to the fact that Naesens et al. only included patients on a glucocorticoid-free immunosuppressive regimen, whereas 82.2% of our patients were still on glucocorticoids during the first year after transplantation. Glucocorticoids are known to increase tacrolimus clearance and thereby decrease the C/D ratio [23]. The retrospective design of our study limits our ability to detect associations between tacrolimus exposure parameters and clinical outcomes, primarily due to insufficient detailed data on tacrolimus levels and corresponding doses, because this study was not designed to detect such a relationship. This highlights the importance of accurate and comprehensive monitoring of tacrolimus levels and dosages in future research to allow for meaningful analysis. Nevertheless, data in both pediatric and adult kidney transplant recipients suggest that higher TacIPV is associated with adverse graft outcomes [12, 24], while the association of a lower C/D ratio with outcome has only been studied in adult kidney transplant recipients [20, 21, 25].

Some limitations must also be noted. The retrospective nature of this registry study prevents causal inferences between age, tacrolimus exposure, and transplant outcomes. In addition, the granularity of our data limited our ability to detail infections such as EBV, BKPyV, and CMV, as they were only recorded in binary form. Despite these challenges, our findings support the exploration of age-specific treatment strategies to optimize outcomes and tailor immunosuppression to the unique needs of different pediatric age groups.

In conclusion, this large registry study of a tacrolimus-based immunosuppressive regimen in European pKTR shows important age-related differences in rejection rates, infection episodes, tacrolimus exposure and clearance. These data suggest that immunosuppressive therapy in pKTR should be tailored according to the age-specific risk profiles of this heterogeneous patient population.

## Acknowledgments/Funding

We acknowledge funding from the Medical Faculty of Heidelberg to Maral Baghai Arassi. The authors gratefully acknowledge the funding of the CERTAIN registry by a grant from the Dietmar Hopp Foundation, the European Society for Pediatric Nephrology (ESPN), the German Society for Pediatric Nephrology (GPN), and by grants from the pharmaceutical companies Astellas and Novartis.

## Statements and Declarations

Burkhard Tönshoff has received research grants from Novartis, Astellas, and Chiesi, and consulting fees from Bristol-Myers Squibb, CSL Behring Biotherapies for Life, Vifor and Chiesi. The other authors declare no conflicts of interest.

## Ethics approval

The study was approved by the Ethics Committee of the Medical Faculty of Heidelberg University and performed in accordance with the World Medical Association Declaration of Helsinki Ethical Principles in the currently valid version.

## Acknowledgments

The authors would like to thank their study nurse Annette Mechler for her continuous and excellent contribution to the CERTAIN registry as well as all contributing CERTAIN study centers with special thanks to Demet Alaygut, Gema Ariceta, Martin Bald, Ortraud Beringer, Laszlo Berta, Antonia Bouts, Salim Caliskan, Henry Fehrenbach, Matthias Hansen, Michael Kaabak, Nele Kanzelmeyer, Günter Klaus, Stephen Marks, Dominik Müller, Hulya Nalcacioglu, Martin Pohl, Michael Pohl, Nikoleta Printza, Alexandr Rumyantsev, Anne-Laure Sellier-Leclerc, Thomas Simon, Oguz Söylemezoglu, Hagen Staude, and Marcus Weitz.

## Data availability statement

The data that support the findings of this study are available from the corresponding author upon reasonable request.

## Abbreviations

ABMR: Antibody-mediated rejection
BKPyV: BK polyomavirus
BSA: Body surface area
BW: Body weight
C/D ratio: Concentration-to-dose ratio
CAKUT: Congenital anomalies of the kidney and urinary tract
CERTAIN: Cooperative European Paediatric Renal Transplant Initiative
CI: Confidence interval
CMV: Cytomegalovirus
CV: Coefficient of variation
EBV: Epstein-Barr virus
eGFR: Estimated glomerular filtration rate
HIV: Human immunodeficiency virus
HR: Hazard ratio
MAD: Mean absolute deviation
MMF: Mycophenolate mofetil
NAPRTCS: North American Pediatric Renal Trials and Collaborative Studies
TacIPV: Tacrolimus intrapatient variability
TCMR: T cell-mediated rejection

## Supplementary Tables

**Table S1:**
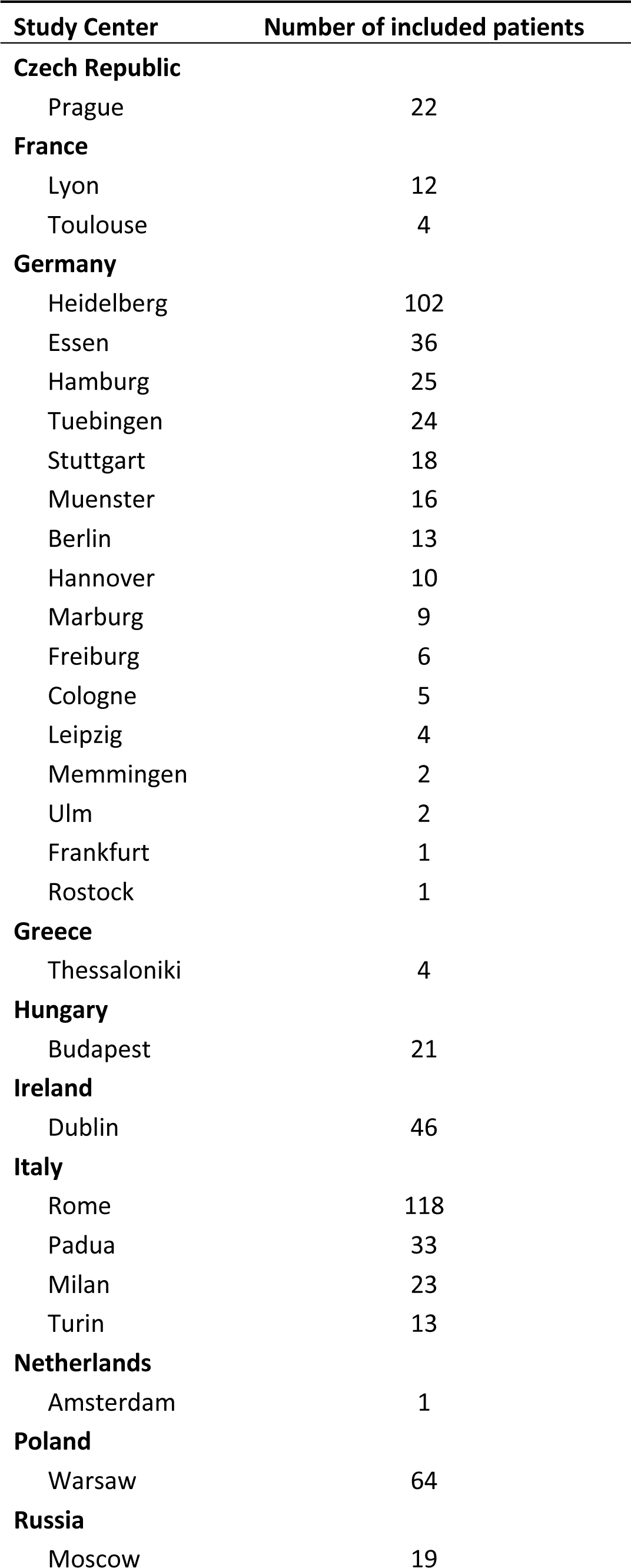
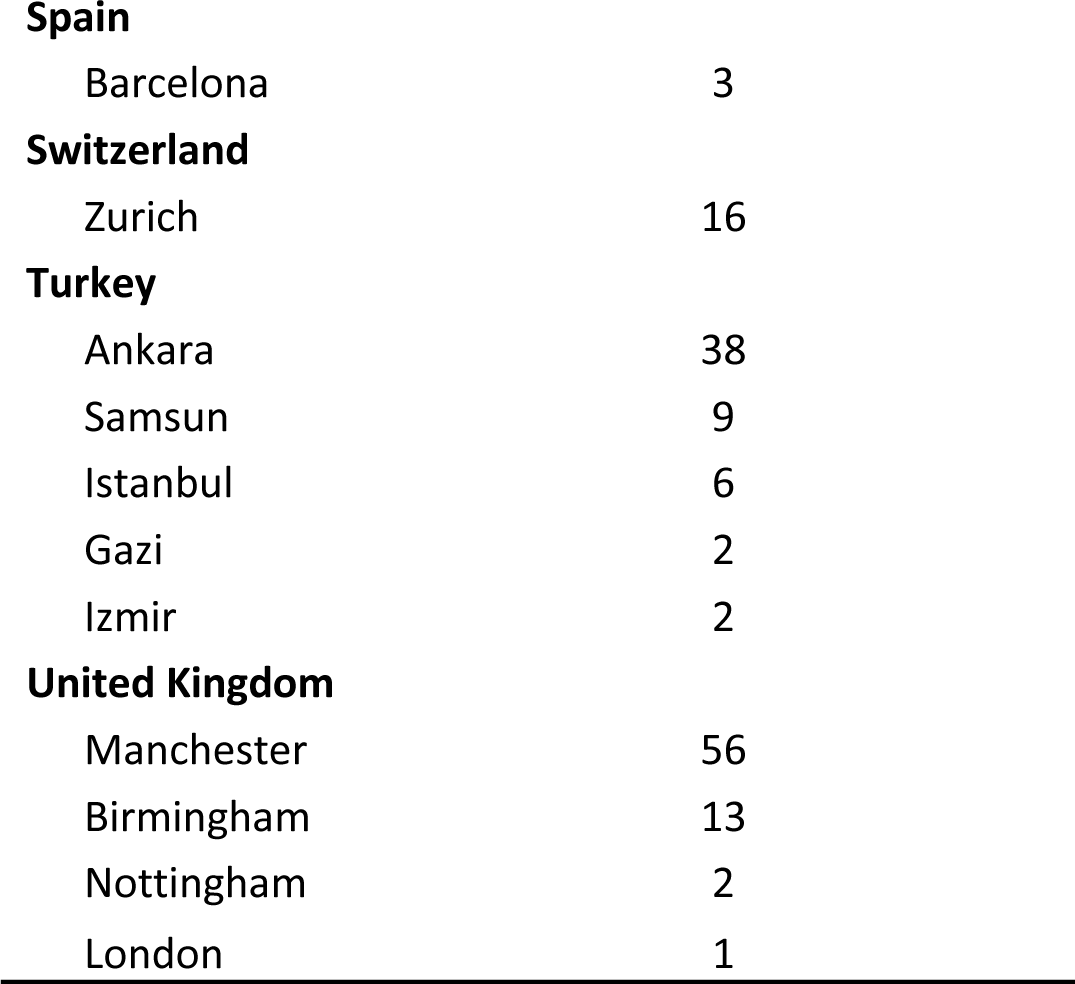
Number of included patients per center.

| **Study Center** | **Number of included patients** |
| --- | --- |
| **Czech Republic** |  |
| Prague | 22 |
| **France** |  |
| Lyon | 12 |
| Toulouse | 4 |
| **Germany** |  |
| Heidelberg | 102 |
| Essen | 36 |
| Hamburg | 25 |
| Tuebingen | 24 |
| Stuttgart | 18 |
| Muenster | 16 |
| Berlin | 13 |
| Hannover | 10 |
| Marburg | 9 |
| Freiburg | 6 |
| Cologne | 5 |
| Leipzig | 4 |
| Memmingen | 2 |
| Ulm | 2 |
| Frankfurt | 1 |
| Rostock | 1 |
| **Greece** |  |
| Thessaloniki | 4 |
| **Hungary** |  |
| Budapest | 21 |
| **Ireland** |  |
| Dublin | 46 |
| **Italy** |  |
| Rome | 118 |
| Padua | 33 |
| Milan | 23 |
| Turin | 13 |
| **Netherlands** |  |
| Amsterdam | 1 |
| **Poland** |  |
| Warsaw | 64 |
| **Russia** |  |
| Moscow | 19 |
| **Spain** |  |
| Barcelona | 3 |
| **Switzerland** |  |
| Zurich | 16 |
| **Turkey** |  |
| Ankara | 38 |
| Samsun | 9 |
| Istanbul | 6 |
| Gazi | 2 |
| Izmir | 2 |
| **United Kingdom** |  |
| Manchester | 56 |
| Birmingham | 13 |
| Nottingham | 2 |
| London | 1 |

**Table S2:** Kidney allograft histopathology graded according to Banff.

| **Graft Histopathology** | **Year 1 post-transplant** |  |  | **Year 2 post-transplant** |  |  |
| --- | --- | --- | --- | --- | --- | --- |
|  | **< 6 years** | **6-12 years** | **> 12 years** | **< 6 years** | **6-12 years** | **> 12 years** |
| Borderline | 19 (9.2%) | 20 (7.2%) | 38 (11.9%) | 7 (3.4%) | 12 (4.3%) | 19 (6.0%) |
| Banff I-III TCMR | 4 (1.9%) | 11 (4.0%) | 25 (7.9%) | 3 (1.5%) | 11 (4.0%) | 7 (2.2%) |
| Chronic TCMR | 0 (0%) | 1 (0.4%) | 1 (0.3%) | 0 (0%) | 0 (0%) | 0 (0%) |
| Mixed ABMR/TCMR | 3 (1.5%) | 5 (1.8%) | 6 (1.9%) | 3 (1.5%) | 1 (0.4%) | 4 (1.3%) |
| Acute ABMR | 5 (2.4%) | 6 (1.8%) | 7 (2.2%) | 2 (1.0%) | 3 (1.1%) | 2 (0.6%) |
| Chronic ABMR | 0 (0%) | 1 (0.4%) | 2 (0.6%) | 0 (0%) | 0 (0%) | 1 (0.3%) |
| BKPyV nephropathy | 2 (1.0%) | 1 (0.4%) | 0 (0%) | 2 (1.0%) | 2 (0.7%) | 2 (0.6%) |
| Sum BPAR | 31 (15.0%) | 42 (15.1%) | 76 (23.9%) | 15 (7.28%) | 27 (9.71%) | 32 (10.1%) |
| Sum BPR | 31 (15.0%) | 44 (15.8%) | 79 (24.8%) | 15 (7.28%) | 27 (9.71%) | 33 (10.4%) |

*TCMR*, T cell-mediated rejection; *ABMR*, Antibody-mediated rejection; *BPAR*, Biopsy proven acute rejections (incl. Borderline); *BPR*, Biopsy proven rejections

**Table S3:** Infection subtypes in the first 2 years post-transplant.

| **Infection subtype** | **< 6 years** | **6-12 years** | **> 12 years** |
| --- | --- | --- | --- |
| Gastroenteritis | 83 (40.3%) | 68 (24.5%) | 48 (15.1%) |
| Gastritis | 5 (2.43%) | 9 (3.24%) | 8 (2.52%) |
| Colitis | 3 (1.46%) | 8 (2.88%) | 5 (1.57%) |
| Peritonitis | 1 (0.49%) | 1 (0.36%) | 0 (0.00%) |
| Lower respiratory infection | 52 (25.2%) | 37 (13.3%) | 26 (8.18%) |
| Pyelonephritis | 50 (24.3%) | 40 (14.4%) | 35 (11.0%) |
| Cystitis | 38 (18.5%) | 51 (18.4%) | 46 (14.5%) |
| BKPyV | 43 (20.9%) | 37 (13.3%) | 30 (9.4%) |
| CMV | 36 (17.5%) | 39 (14.0%) | 49 (15.4%) |
| EBV | 65 (31.6%) | 48 (17.3%) | 32 (10.1%) |
| Sepsis | 26 (12.6%) | 19 (6.8%) | 16 (5.03%) |
| Hepatitis | 2 (0.97%) | 3 (1.08%) | 2 (0.62%) |
| HIV | 0 (0.00%) | 1 (0.36%) | 0 (0.00%) |
| Pneumocystis jirovecii | 2 (0.97%) | 0 (0.00%) | 1 (0.31%) |
| Other fungal infection | 9 (4.37%) | 7 (2.52%) | 9 (2.83%) |
| Other viral infection | 76 (36.9%) | 62 (22.3%) | 53 (16.7%) |

BKPyV, BK polyomavirus; CMV, Cytomegalovirus; EBV, Epstein-Barr virus; HIV, Human immunodeficiency virus

## Supplementary Figures

**Fig. S1:**
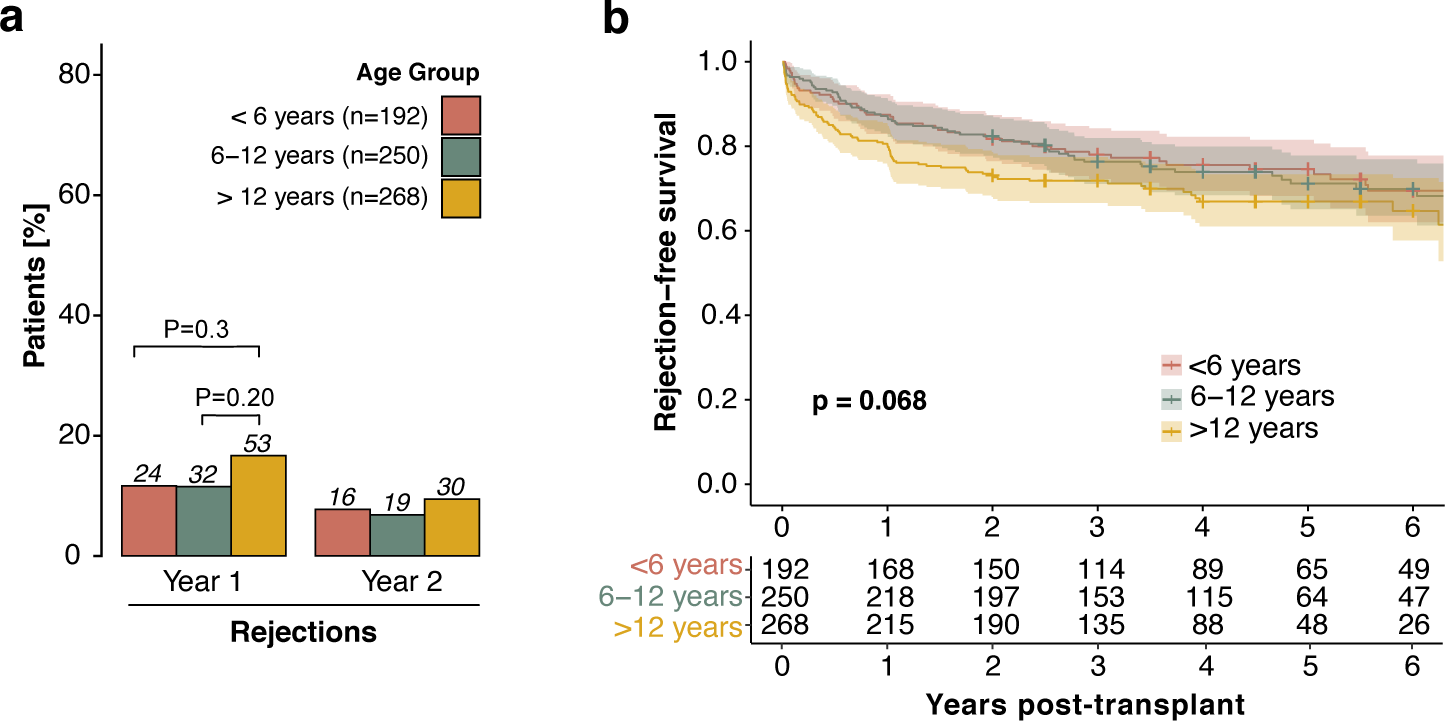
Biopsy-proven rejection episodes in patients without previous kidney transplants stratified by age group. (a) Incidence of rejection in the first two years post-transplant, stratified by age group and year post-transplant. The first event of each patient in each year is considered for analysis. Benjamini-Hochberg adjusted P values are reported. **(b)** Rejection-free survival: Kaplan-Meier curves illustrating the probability of rejection-free survival over the entire follow-up period post-transplant, with age groups distinguished by color coding. Log-rank P values from the Kaplan-Meier analysis are provided to indicate the statistical significance of differences between age groups.

## Supplementary Methods

### Description of the CERTAIN registry: completeness and quality of data

CERTAIN provides a detailed data collection that allows an in-depth characterization of specific patient cohorts. Data are collected prior to kidney transplantation, at 1, 3, 6, 9, and 12 months post-transplant, and at 6-month intervals thereafter. In addition, the CERTAIN dataset allows for detailed and flexible documentation of the post-transplant follow-up through continuous entry of any number of relevant data (e.g., laboratory values, drug therapy). Specific case report forms (CRFs) collect detailed and accurate information on relevant data and events of pediatric kidney transplantation in the peri- and post-transplant course. There are two datasets, the minimum required dataset and the extended dataset. The minimum required dataset is mandatory for all participating centers. The extended data set provides a deeper insight into the clinical course and treatment of patients by documenting additional items, some of which are predefined and some of which can be defined by the participating center. The CERTAIN web application (accessible via http://www.certain-registry.eu/RegApp) has an automatic and manual data validation functionality. During data entry, the data record is automatically validated against predefined plausibility ranges. In addition, the system has an integrated manual quality assurance process. First, documented data must be approved locally at the site; second, a data quality manager at the registry headquarters randomly checks data for plausibility. Only data that pass this quality assurance process are added to the research database. These functions are available anytime, anywhere and require only a standard Web browser and Internet access.

## Notes

### Competing Interest Statement

Burkhard Tonshoff has received research grants from Novartis, Astellas, and Chiesi, and consulting fees from Bristol-Myers Squibb, CSL Behring Biotherapies for Life, Vifor and Chiesi. The other authors declare no conflicts of interest.

